# Center-of-Mass Work Organization Supplements Walking Speed: a Biomechanical Characterization of Hemiparetic Gait

**DOI:** 10.64898/2026.03.12.26348298

**Authors:** Seyed-Saleh Hosseini-Yazdi, Karson Fitzsimons, John EA Bertram

## Abstract

**Background and Purpose:** Walking speed is the dominant clinical metric used to classify post-stroke hemiparetic gait severity. However, speed does not describe how mechanical energy is generated and redistributed. We tested whether whole-body center-of-mass (COM) work patterns provide a biomechanically grounded supplement to speed-based severity classification.

**Methods:** Limb-specific COM power and work were computed from ground reaction forces using the individual-limbs method across five walking speeds (0.2–0.7 m/s). We quantified net COM work index of asymmetry (IA_Wnet), positive COM work asymmetry (IA_Wpos), and the Propulsion–Support Ratio (PSR = impFy/impFz). Piecewise and quadratic regressions were used to assess speed-dependent trends.

**Results:** IA_Wnet remained elevated across speeds and showed no significant high-speed association. IA_Wpos demonstrated a significant quadratic relationship with speed (p=0.023, R²=0.23), decreasing near 0.5 m/s before rising again. Paretic limb PSR remained constrained and exhibited a quadratic association (p=0.012, R²=0.14), while unaffected limb PSR declined significantly at higher speeds (p=0.019, R²=0.38). Below 0.5 m/s, COM power profiles collapsed to a two-phase pattern without paretic limb push-off; at ≥0.5 m/s, a four-phase structure emerged.

**Conclusion:** Increasing walking speed did not normalize interlimb mechanical imbalance. COM work organization revealed a biomechanical transition near 0.5 m/s and distinguished compensation from recovery-based restoration. Supplementing speed with COM work and propulsion–support metrics may refine severity stratification and guide mechanism-targeted rehabilitation.

## Introduction

Stroke remains a leading cause of long-term adult disability, and impaired walking is among its most prevalent and persistent sequelae. Walking dysfunction has been reported in a large majority of stroke survivors, and residual gait impairment frequently persists despite rehabilitation, limiting community participation and independence ^1^. The characteristic post-stroke hemiparetic gait reflects a coupled impairment of balance control and step-to-step redirection of the body’s center of mass (COM), these constraints give rise to heterogeneous patterns of spatiotemporal and kinetic asymmetry ^1^.

In clinical practice and clinical trials, walking speed is the most common single measure used to stratify ambulatory status, track progress, and inform clinical rehabilitation strategies. Speed-based functional ambulation categories (e.g., household, limited community, and community ambulation) have a long history in stroke rehabilitation research and are widely used because they are simple, reliable, and associated with functional outcomes ^2^. Contemporary stroke rehabilitation guidance continues to recommend standardized capacity-based assessments such as the 10-m walk test (gait speed) and the 6-minute walk test (walking endurance) as core measures of walking recovery ^3^.

However, speed is fundamentally an outcome rather than a mechanistic descriptor of how walking is produced. Two individuals may walk at similar speeds yet rely on very different biomechanical strategies, degrees of compensation, and margins of stability ^4,5^. This limitation is particularly salient in hemiparetic gait, where improved speed can arise from (i) recovery-based restoration of paretic limb function, or (ii) compensatory redistribution of mechanical demands to the non-paretic limb and proximal joints ^6^.

Because hemiparetic walking is visibly asymmetric, research and rehabilitation studies frequently quantify step length asymmetry, step time/double-support asymmetry, and related spatiotemporal descriptors ^6^. Yet spatiotemporal asymmetries are widely recognized as mixed markers that may reflect both impairment and compensation. For example, step length asymmetry has been shown to represent compensatory mechanisms and is associated with underlying kinetic deficits (e.g., plantarflexor moment impulse reductions) rather than serving as a direct readout of the causal impairment itself ^6^. Moreover, the relationship between reducing asymmetry and improving energetic performance is heterogeneous and depends on the direction and structure of asymmetry and baseline impairment ^7^.

In parallel, stroke biomechanics has emphasized limb-level propulsion metrics—particularly reduced paretic limb anterior–posterior ground reaction force and propulsive impulse—because paretic limb propulsion is strongly linked to walking performance and is a frequent target of intervention studies ^8^. While informative, these limb-level metrics still leave a key gap: they do not directly characterize how the gait system regulates whole-body COM energy and redirection, nor do they provide a unified way to interpret the broad constellation of observed asymmetries as a coherent severity phenotype.

A complementary and more integrative framework comes from the mechanics and energetics of walking as a COM modulation problem. In dynamic-walking theory, the step-to-step transition requires mechanical work to redirect the COM velocity vector from one pendular arc to the next^9,10^; this transition work is a major determinant of metabolic cost ^11^ and is tightly coupled to the key features of the step-to-step transition: coordination of leading-limb collision and trailing-limb push-off ^10^. Importantly, standard “combined-limbs” external work estimates can underestimate work during double support because simultaneous positive and negative work by the trailing and leading limbs can cancel in the summed signal; the individual-limbs COM power/work method resolves this and provides limb-specific COM work contributions during the critical transition phase ^9^.

Applied to post-stroke hemiparetic walking, individual-limb and COM-level analyses have repeatedly highlighted a characteristic reduction in paretic limb push-off work and altered distribution/timing of mechanical work across the limbs, with consequences for overall work demands and energetic cost ^12^. For example, studies using individual-limb mechanical analyses in chronic stroke have compared mechanical asymmetries across the step-to-step transition and single support and related these asymmetries to impairment strata defined by gait speed ^13^. Studies that explicitly revisits hemiparetic walking from individual-limb COM work to joint-level work further support the view that increased mechanical work—rather than reduced “efficiency” alone—contributes to the elevated metabolic cost following stroke, emphasizing the importance of identifying how mechanical work is generated and redistributed ^12^. Additionally, COM-directed propulsive forces and their timing have been linked to metabolic energetics post-stroke, reinforcing the relevance of COM-level descriptors beyond traditional spatiotemporal metrics ^14^.

Despite these mechanistic foundations, COM work patterns are rarely used as the primary construct to categorize severity in post-stroke hemiparetic gait from a biomechanical viewpoint. Instead, severity is typically inferred from walking speed categories or from observable asymmetries in step parameters, with COM work most often appearing—if at all—as a specialized research analysis rather than a severity taxonomy ^2^. This mismatch is notable because many of the hallmark asymmetries of hemiparetic gait can be interpreted as downstream consequences of constrained COM work capacity—particularly constraints imposed by impaired paretic limb work and the resulting limited ability to express a timely, appropriately phased positive work pattern during stance and transition. From this view, step length asymmetry, double-support prolongation, and propulsion redistribution are not independent deficits but coordinated adaptations to maintain forward progression and avoid instability when the COM work pattern is compromised ^6,15^ (Figure 1).

**Figure 1:**
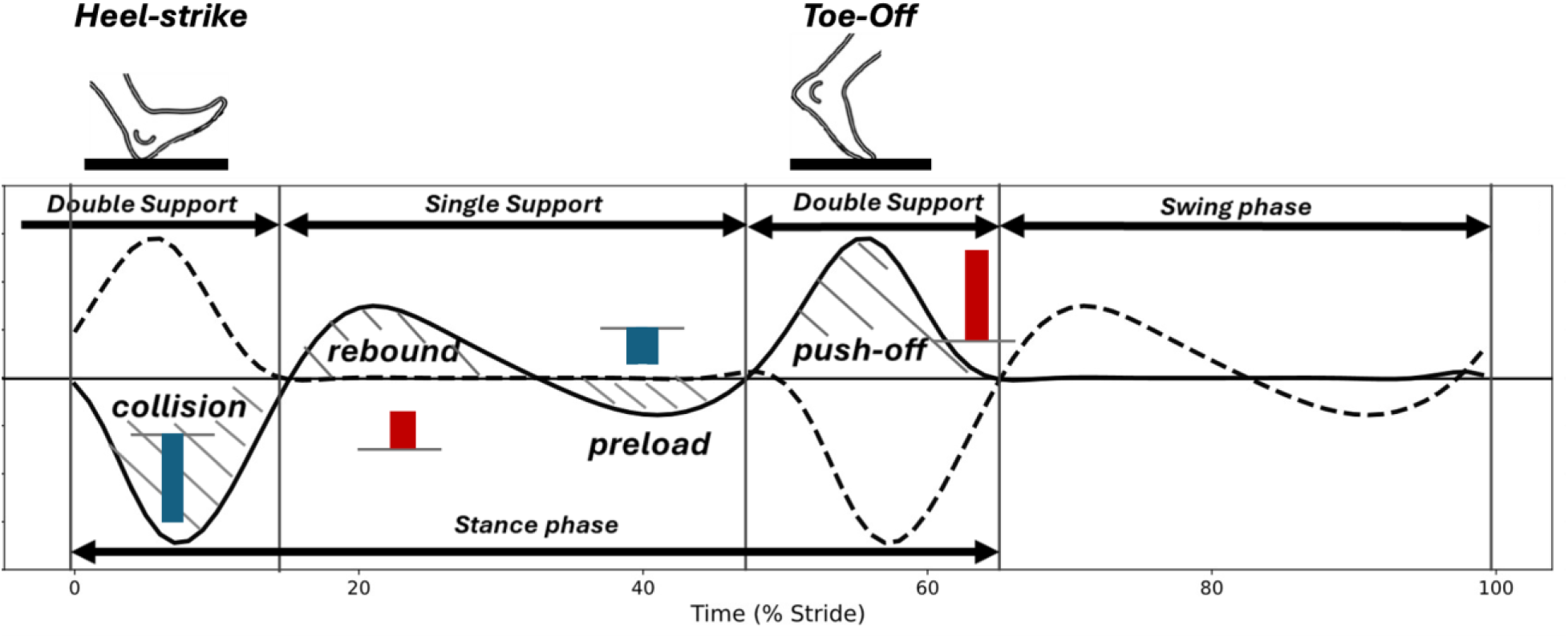
It is shown that in healthy nominal walking, the center of mass (COM) power depicts four distinct phases, starting with heel strike collision negative work. During the single support, the positive and negative work follow which end to push-off (positive work) in late stance (adopted from ^16^). The associated work of each phase is calculated by time integration of the associated power profile. Negative work is indicated by dark blue columns while positive work are indicated by dark red columns.

Accordingly, there is a strong rationale to move beyond speed and step-parameter asymmetry as dominant descriptors and to test whether COM work patterns can serve as a biomechanically grounded indicator of hemiparetic gait severity. Building on dynamic walking theory and individual-limb COM power methodology ^10,11^, we propose that severity can be characterized biomechanically by how effectively the paretic and non-paretic limbs (i) support and redirect the COM through stance and (ii) coordinate positive and negative work during the step-to-step transition and subsequent single support ^17^. Such a categorization has the potential to (1) explain why diverse asymmetries emerge, (2) discriminate compensation from recovery-based restoration at similar speeds, and (3) provide mechanistic targets aligned with known post-stroke deficits in push-off and transition dynamics.

## Materials and Method

Since the COM energy is constantly modulated by work generation^18^, we need to quantify the COM work through the ground reaction forces (GRF) of walking. GRFs are low pass filtered (cut-off = 10Hz, third order) with no phase change and utilized to calculate the COM velocity ^11^. To remove drift, the COM speed components are high pass filtered (cutoff = 0.25 Hz, third order) ^19^. Accordingly, the limb specific COM power ^9,11,15^ is calculated as *P*_*limb*_ = *v⃗*_*COM*_. *F⃗*_*COM*_. Based on the profile of the COM power, if a distinct four phase work pattern is present (Fig. 1), we compute work for each phase of stance. If the COM power does not show the conventional profile ^15^, we limit work computation to total step positive and negative work only.

Accordingly, we develop some metrics to assess the asymmetries of hemiparetic walking based on COM work and step impulses in the anterior-posterior (PA) and vertical directions. We compare the metrics with COM work distributions and against each other to examine which approach provides the clearest perspective regarding biomechanical evaluation of hemiparetic walking.

The first metric we introduce is Net-work asymmetry that quantifies the imbalance in net COM energy exchange between limbs and reveals whether one limb persistently supplies energy while the other dissipates it. Net-work asymmetry as speed changes will indicate that, despite changes in propulsion sharing, hemiparetic walking remains energetically compensated for rather than mechanically restored.

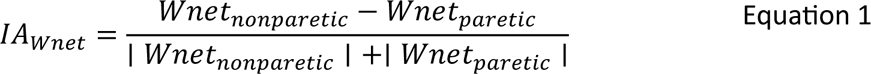

We expect *IA*_*Wnet*_ > 0 for hemi-paretic walking. In other words, we anticipate the unaffected side to compensate for the energy output deficit of the paretic-side.

Next, we define the propulsion index of asymmetry for positive work generation as a percentage asymmetry about the mean, defined by:

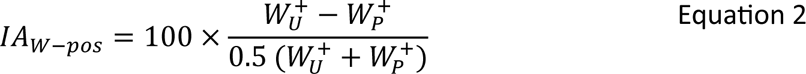

In this formulation, *W_U_*^+^ and *W_P_*^+^ are the positive COM work per step performed by the unaffected and paretic limbs, respectively. This quantity represents how much mechanical energy each limb adds to the COM, primarily during push-off, or when mechanics are disrupted, sometimes during rebound. Importantly, this asymmetry index does not measure how much work each limb performs in absolute terms given by *W*^+^, nor how efficiently stance is converted into propulsion. Instead, it measures how far the system is from symmetric propulsion, i.e., how unequal the limbs are relative to the mean propulsive contribution. For this reason, it is best understood as a symmetrical metric rather than a mechanistic metric. This distinction is critical for interpreting results. In severe cases when walking speed is low, we expect small paretic limb positive work while the unaffected limb work must be large, driving the index toward its maximum, indicating extreme asymmetry. As severity declines and walking speed increases, paretic positive work must rise while unaffected limb work ought to decrease, making *W_U_*^+^ + *W_P_*^+^ more balanced and causing the index to drop. Thus, the reduction of this index with speed must provide a clear quantitative indicator that propulsion becomes progressively shared between limbs as paretic push-off emerges. Hence, we expect that this metric captures an improvement in step-to-step mechanical coordination that is not directly visible from timing measures alone.

Our final indicator is the Propulsion–Support Ratio (PSR) that is defined as the anterior–posterior impulse normalized by vertical impulse during stance:

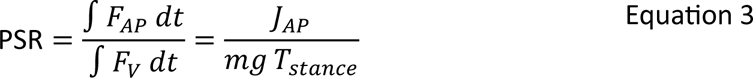

PSR therefore quantifies how effectively a limb converts vertical load bearing during stance into forward momentum of the center of mass. Unlike COM work metrics, PSR is a momentum-based descriptor derived directly from ground reaction forces and is insensitive to the timing of propulsion, making it particularly suitable for identifying propulsion inefficiency in hemiparetic gait where the limb supports body weight but fails to generate adequate forward progression.

### Participants

Eligible participants were identified by physiotherapists and clinicians working in the Neuro Rehab Unit of the Foothills Hospital in Calgary, Alberta, Canada. Participants were included if they were over the age of 18 and had experienced their first ischemic or hemorrhagic stroke within the past 6 months. Participants were excluded if they had other history of significant neurological injury or disease, lower limb orthopedic issues, excessive pain or comorbidities that prevented them from completing walking tasks, and if they had experienced a cerebellar stroke. Participants were recruited as part of a larger study on the impacts of split-belt treadmill walking during rehabilitation, with written informed consent obtained according to a protocol approved by the Conjoint Health Research Ethics Board at the University of Calgary (Ethics ID REB21-1576).

Eleven participants (5 female; 6 male) undergoing post-stroke inpatient rehabilitation were recruited. The mean (SD) age was 54.6 (11.5) years, the mean (SD) time post-stroke at first assessment was 10.0 (7.0) weeks.

### Assessment Protocol

Before the first session of treadmill walking, each participant’s overground self-selected walking speed was measured as the average of two 10m walk tests. Participant endurance was evaluated using a 6 minute walk test and the Fugl-Meyer Lower Extremity assessment was conducted as a clinical baseline. Participants were permitted the use of gait aids, including canes and walkers, when completing the overground walking tests.

At each session, participants completed at least 2 minutes of walking on a on a split belt instrumented treadmill (Bertec Corp, Columbus Ohio) with embedded force plates recording ground reaction force (GRF) at 1000Hz. Each participant wore a harness connected to patient lift capable of fall arrest and were allowed hand contact with a handrail located at the front of the treadmill. A semi-immersive virtual reality pathway was displayed across screens in front of the treadmill, with the visual flow of the pathway matched to the speed of the treadmill. As an additional precaution, heart rate and blood pressure was measured before and after each walking bout. These sessions were completed as part of each participants prescribed rehabilitation under the direct supervision of their clinical physiotherapist.

Each participant completed a variable number of sessions over the course of their inpatient stay, with up to 3 sessions per week. Treadmill speed was increased over the course of the sessions at the discretion of the presiding physiotherapists, reflecting the improvements in functional ability of the participants.

## Results

To determine whether whole-body COM work patterns provide a mechanistically grounded descriptor of severity of impairment in hemiparetic walking, we quantified limb-specific COM work distributions and derived asymmetry and propulsion–support indices across five walking speeds (0.2–0.7 m. s^−1^). We first characterized qualitative transitions in vertical and anterior–posterior ground reaction forces and COM power profiles, followed by quantitative evaluation of (i) net COM work asymmetry (AI_W_net_), (ii) positive COM work asymmetry (AI_W_pos_), and (iii) the Propulsion–Support Ratio (PSR). Regression analyses were performed to evaluate whether these indices varied systematically with walking speed and to identify potential transition points in mechanical function and organization.

We categorized the subjects’ results based on their walking speed into five bins: 0.2, 0.3, 0.4, 0.5 and 0.7 m. s^−1^. The vertical ground reaction forces did not show the typical double peak profile of normal walking up to 0.5 m. s^−1^. The paretic side vertical force peaks are also lower than the unaffected side. For PA force, the paretic side performed larger breaking impulse while the unaffected side performed larger propulsive impulse. For the COM power, the four-phase power profile was not observed up to 0.5 m. s^−1^. Above 0.5 m/s the paretic side was able to perform some push-off (Figure 2).

**Figure 2:**
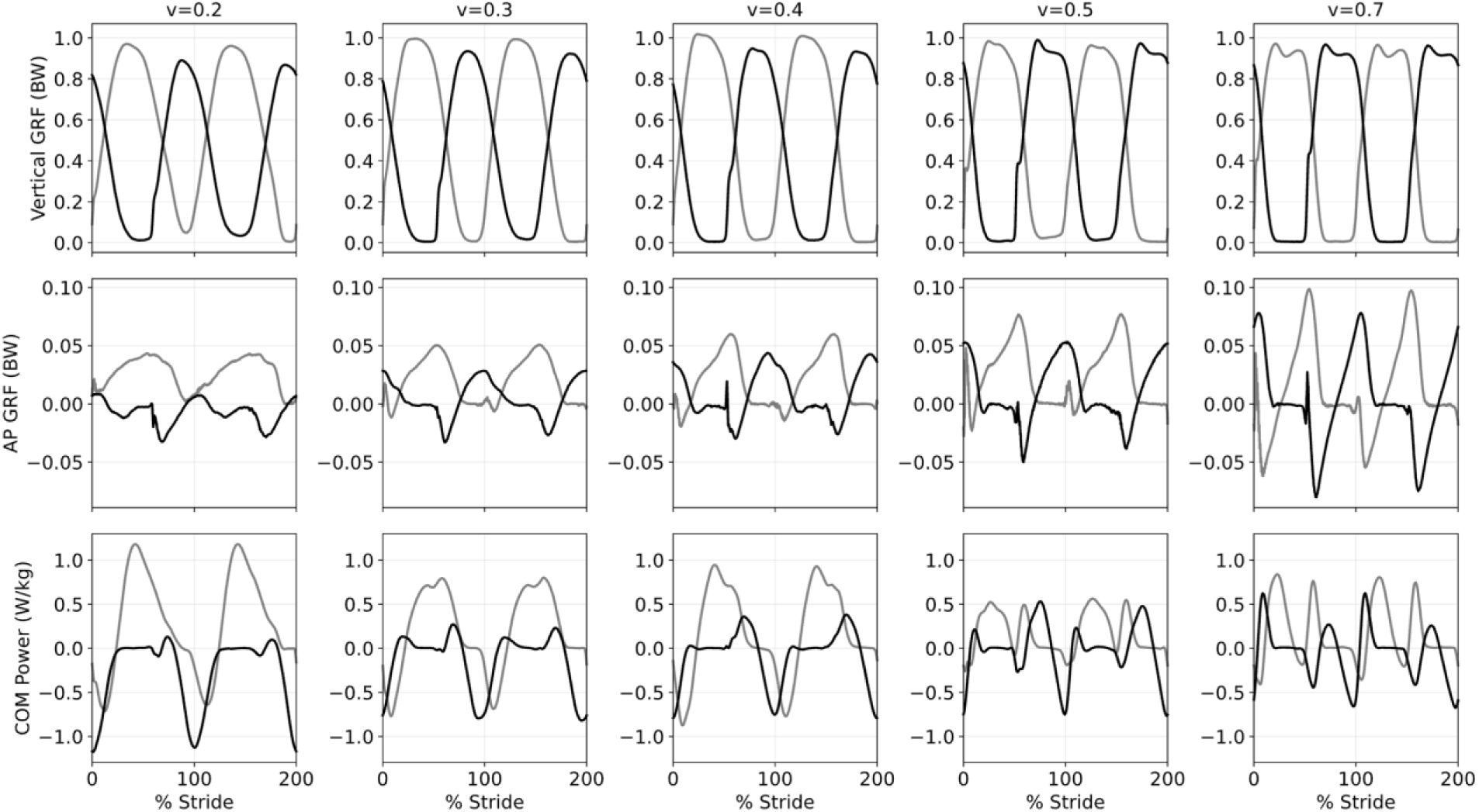
The hemi-paretic walking data collection based on walking speed: 0.2, 0.3, 0.4, 0.5 and 0.7 *m*. *s*^−1^. The top row shows the average vertical ground reaction force, the middle row is for the average PA ground reaction force, and the bottom row is for the average COM power. For speeds less than 0.5 *m*. *s*^−1^, the COM power profile shows a two-phase trajectory. The four -phase trajectory emerges for walking speeds ≥ 0.5 *m*. *s*^−1^. The paretic traces are shown in black while unaffected sides are depicted in gray.

Since we could not recognize the four-phase power profile for speeds less than v =0.5 m. s^−1^, we present only the positive and negative COM work sequence when there was no visible push-off work on either limb. In other words, when there was no distinguishable push-off the COM power profile was reduced to two phases: positive and negative work while the orders of paretic and unaffected sides were inverse of each other (Figure 3). When push-off appeared in late stance *v* =0.5 and 0.7m. s^−1^), the four-phase COM power profile emerged, and the magnitude of COM work declined materially.

**Figure 3:**
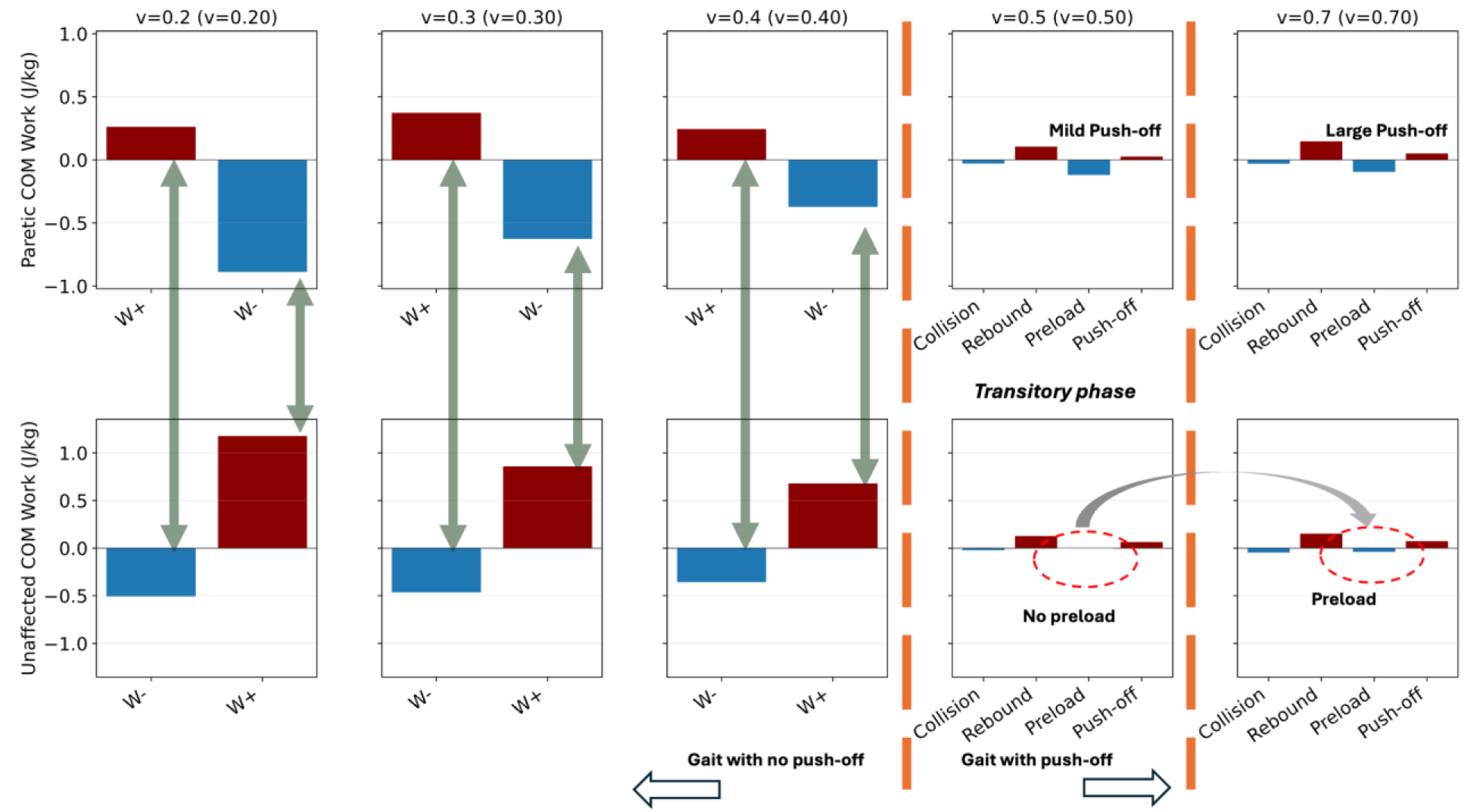
The COM work components of hemi-paretic walking: the top row indicates the paretic side while the bottom row represents the unaffected side. We present the positive work as dark red whereas blue indicates step negative work. For *v* =0.2, 0.3, and 0.4 *m*. *s*^−1^ where the push-off is not detected, there are only two dominant work phases: positive and negative. At *v* =0.5 *m*. *s*^−1^ paretic push-off appears and both sides show four-phase COM power profiles. At *v* =0.5 *m*. *s*^−1^, single support preload is negligible, yet its magnitude rises at *v* =0.7 *m*. *s*^−1^.

Net COM work asymmetry (IA_W_net_; Unaffected (U) vs Paretic (P)) remained elevated across all speeds, indicating persistent whole-body mechanical imbalance between limbs. At 0.2 m. s^−1^, asymmetry was 0.783 ± 0.582 and increased to 0.846 ± 0.489 at 0.3 m. s^−1^. At 0.4 m. s^−1^, asymmetry reached a maximal value (1.000 ± 0.174), indicating complete dominance of net work by the unaffected limb. Although asymmetry decreased at 0.5 m. s^−1^ (0.684 ± 0.428), it increased again at 0.7 m. s^−1^ (0.822 ± 0.262). Importantly, net work asymmetry did not decrease monotonically with speed, suggesting that increasing walking velocity did not normalize interlimb mechanical contribution. Instead, whole-body work remained predominantly redistributed toward the unaffected limb across speeds.

Hence, to determine whether net COM work asymmetry varied systematically with walking speed, a piecewise linear regression was performed separating slower and higher speed ranges based on maximizing the R^2^ and significance of the proposed fit. In the slower speed range (<-0.5 m. s^−1^), net work asymmetry demonstrated a positive slope (slope = 1.28), approaching statistical significance (p = 0.053) with modest explanatory power (R² = 0.10). This trend indicates that, at lower walking speeds, increases in speed were associated with a tendency toward greater interlimb net work imbalance, although the relationship did not reach conventional statistical significance with this relatively small sample size. In contrast, within the higher speed range (>0.5 m. s^−1^), the slope was smaller (slope = 0.69), non-significant (p = 0.48), and explained minimal variance (R² = 0.04), indicating no meaningful association between walking speed and net COM work asymmetry at higher speeds.

Together, these findings suggest that net COM work asymmetry is largely insensitive to increases in walking speed, particularly beyond the slow-speed domain. The modest trend observed at slower speeds does not persist at higher speeds, reinforcing the interpretation that interlimb mechanical imbalance is not normalized by increasing walking velocity. Rather, work and power asymmetry remains a persistent feature of hemiparetic walking across speeds. The point of break suggests a transition in asymmetrical index about *v* =0.5 m. s^−1^ (Figure 4A). The positive COM work asymmetry (IA_W_pos_), representing propulsion asymmetry, was markedly elevated at slower speeds and demonstrated a nonlinear pattern with speed. At 0.2 m. s^−1^, propulsion asymmetry was 103.38 ± 66.42%, indicating substantial dominance of the unaffected limb. At 0.3 and 0.4 m. s^−1^, asymmetry remained high (75.54 ± 52.49% and 79.92 ± 44.23%, respectively). A pronounced reduction was observed at 0.5 m. s^−1^ (25.26 ± 28.47%), followed by a partial increase at 0.7 m. s^−1^ (55.24 ± 45.51%). Despite some reduction at intermediate speeds, propulsion asymmetry persisted at all walking conditions, indicating that positive work redistribution was not fully restored even when speed increased.

**Figure 4:**
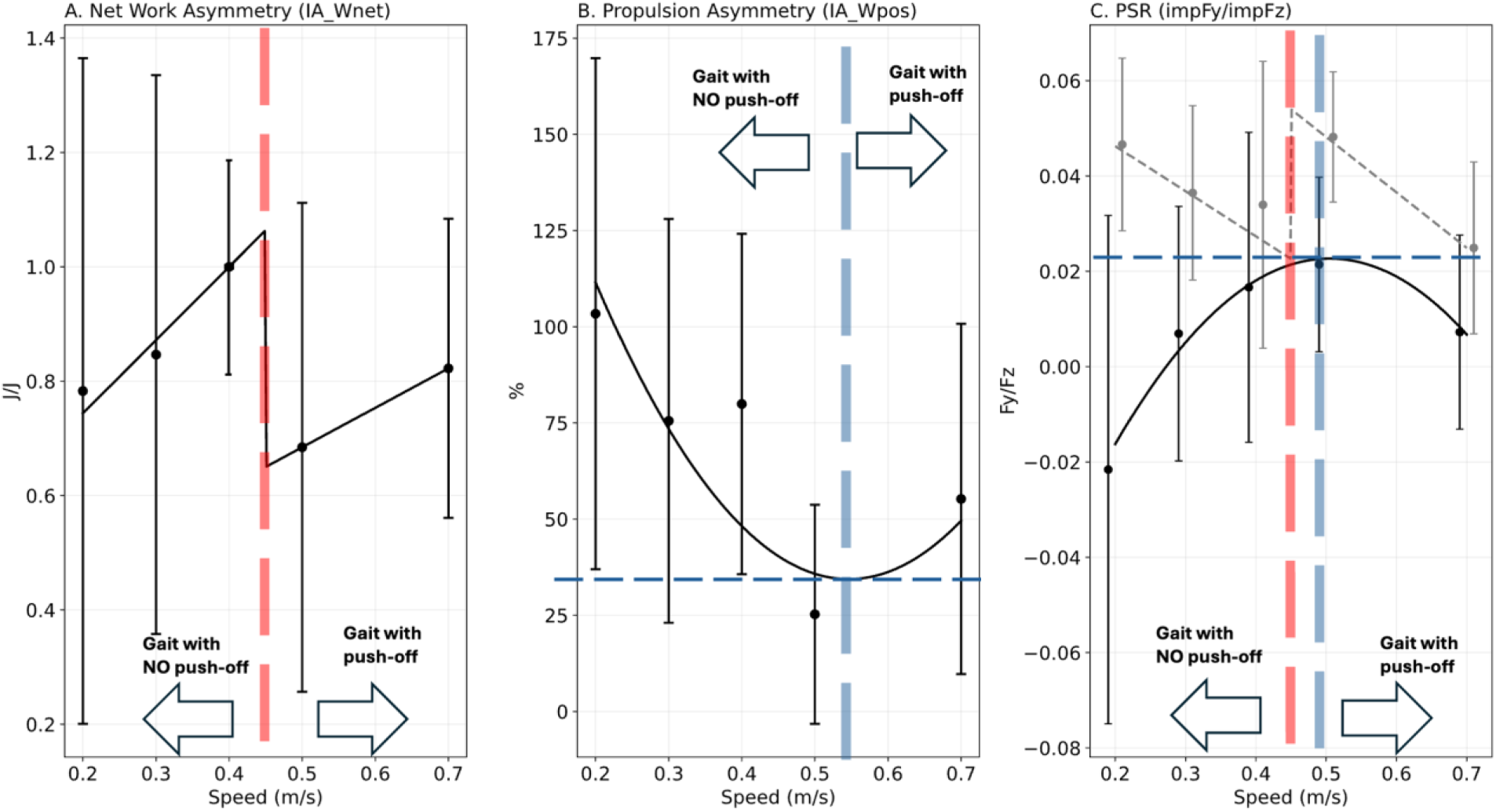
Hemiparetic walking asymmetry evaluation indices: (A) Net work asymmetry index measuring the asymmetry in net work performance of paretic and unaffected side, (B) the Propulsion index of asymmetry representing the asymmetry in positive work of either side, and (C) the PSR index for paretic and unaffected sides. All indices suggest a transition in the state of hemiparetic walking around *v* = 0.5 *m*. *s*^−1^. The solid line represents the PSR for the paretic side while the dashed gray line show the unaffected side. The detected transitions are also indicated by vertical dashed lines.

Given the nonlinear pattern observed across walking speeds, a quadratic regression was performed to evaluate the association between speed and positive COM work asymmetry (IA_W_pos_). The quadratic term was statistically significant (quadratic coefficient (β2) = 642.891, p = 0.023), with the model explaining 23% of the variance (R² = 0.23). This indicates a significant nonlinear relationship between walking speed and propulsion asymmetry. Consistent with the descriptive data, propulsion asymmetry was markedly elevated at slower speeds, decreased at intermediate speeds, and increased again at higher speeds. The significant quadratic term confirms that this U-shaped pattern is not attributable to random variability but reflects a systematic speed-dependent redistribution of positive work between limbs. Importantly, despite partial reduction at intermediate speeds, propulsion asymmetry did not normalize across the tested speed range. These findings suggest that paretic positive work capacity does not scale proportionally with speed demands and that propulsion redistribution remains a defining biomechanical feature of hemiparetic walking. As such, we also found a trend transition at about *v* =0.5 m. s^−1^(Figure 4B).

The paretic limb Propulsion Support Ratio (PSR) showed very low values at slower speeds and remained substantially lower than the unaffected limb across conditions. At 0.2 m. s^−1^, paretic limb PSR was negative (−0.0216 ± 0.0533), indicating an absence of effective forward impulse relative to vertical support. Paretic limb PSR increased slightly with speed (0.0069 ± 0.0267 at 0.3 m. s^−1^; 0.0166 ± 0.0325 at 0.4 m. s^−1^; 0.0214 ± 0.0183 at 0.5 m. s^−1^) but declined again at 0.7 m. s^−1^(0.00724 ± 0.0204). In contrast, unaffected limb PSR remained consistently positive and larger across speeds (0.0466 ± 0.0181 at 0.2 m. s^−1^; 0.0365 ± 0.0183 at 0.3 m. s^−1^; 0.0340 ± 0.0301 at 0.4 m. s^−1^; 0.0482 ± 0.0137 at 0.5 m. s^−1^; 0.0249 ± 0.0180 at 0.7 m. s^−1^). Across all speeds, unaffected limb PSR exceeded paretic limb PSR, confirming that forward impulse generation relative to load support remained disproportionately shifted toward the unaffected limb.

A quadratic regression model was applied to evaluate the relationship between walking speed and paretic limb PSR. The quadratic term was statistically significant (quadratic coefficient (β2) = 0.419, p = 0.012), with the model explaining 14% of the variance (R² = 0.14), indicating a significant nonlinear association between speed and paretic limb propulsion relative to vertical support. This nonlinear relationship reflects modest increases in paretic limb PSR at intermediate speeds, with attenuation at higher speeds. Despite statistical significance, the relatively low R² indicates that speed accounts for only a limited proportion of the variability in paretic propulsion-support coupling. Importantly, absolute paretic limb PSR values remained small across speeds, indicating persistent constraint in forward impulse generation relative to load support capacity.

For the unaffected limb, a piecewise linear model provided the best fit. In the slow-speed domain, the slope was negative but not statistically significant (slope = 0.095, p = 0.131), explaining minimal variance (R² = 0.06). In contrast, in the higher-speed domain, PSR demonstrated a significant negative slope (slope = −0.116, p = 0.019), with substantially greater explanatory power (R² = 0.38). This pattern indicated that at higher walking speeds, unaffected limb propulsion relative to support decreased systematically. The significant negative association in the high-speed range suggested a redistribution or modulation of unaffected limb propulsion as speed increased. Similar to prior indices, PSRs of unaffected and paretic sides indicated a transition at *v* =0.5 m. s^−1^ (Figure 4C).

## Discussion

This study is an attempt to add a mechanistic layer to understand post-stroke gait severity characterization by showing that the whole-body COM work pattern and associated derived metrics, i.e. net work asymmetry, positive work asymmetry, and propulsion–support coupling, contain information that is not captured by walking speed variations alone. Gait speed is rightly central in stroke rehabilitation—endorsed in major practice guidelines and widely used in trials because it is feasible, reliable, and meaningful to function ^3^. However, our results demonstrate that speed-based severity classification can be mechanically incomplete and potentially misleading, because higher speed can coexist with persistent interlimb mechanical imbalance and constrained paretic limb propulsion-support coupling.

Net COM work asymmetry (AI_W_net_) remained high across speeds and showed only weak speed dependence. Piecewise regression suggested a borderline trend in the slow-speed domain (*v*_*ave*_ < 0.5 m. s^−1^), but no meaningful relationship at higher speeds. This indicates that increasing speed did not systematically reduce interlimb net work imbalance, consistent with prior individual-limb analyses showing robust asymmetries in mechanical power after stroke across phases of the stride, with the non-paretic limb producing more positive net mechanical power ^13^. The implication is that net work asymmetry may represent a relatively confounding feature of hemiparetic walking—more reflective of underlying capacity constraints than of speed per se.

In contrast, positive COM work asymmetry (AI_W_pos_) exhibited a significant nonlinear association with speed. This pattern supports the interpretation that propulsion sharing is not a steadily improving function of speed but instead reflects speed-dependent reorganization of how forward progression is achieved. This helps contextualize why step-parameter symmetry changes do not always map onto metabolic or mechanical improvement. Prior study has shown that individuals can achieve more symmetric stepping while still walking with a high energetic demand and persistent kinetic constraints^20^. Related study also highlights the heterogeneity in how asymmetry, training response, and energetic outcomes relate across individuals post-stroke^7^. Taken together, these findings reinforce that COM work distribution provides a clearer indication of the underlying mechanical movement strategy than step symmetry alone.

PSR findings further sharpen the mechanistic interpretation. The paretic limb PSR showed a significant quadratic relationship with speed, yet remained small overall, indicating a persistent limitation in converting stance support to forward impulse. This is consistent with broader stroke and propulsion studies emphasizing paretic limb propulsion deficits as core contributors to the severity of hemiparetic deficits and as important therapeutic targets ^21^. Notably, unaffected-limb PSR was best described by a piecewise model: the slow-speed slope was non-significant (*v*_*ave*_ < 0.5 m. s^−1^), whereas the high-speed slope was significantly negative. This suggests that once speed reaches a higher domain, the non-paretic limb’s propulsion relative to support decreases systematically—consistent with the idea that compensation is not unlimited and may be reallocated as task demands increase.

Major stroke rehabilitation guidance and outcomes literature justifiably centers on gait speed, and many high-impact trials use gait speed as a primary endpoint ^3^. Yet methodologic studies also caution that short-distance speed can overestimate broader locomotor capacity and that measurement choices matter ^22^. Our results extend that caution from ***capacity*** to ***mechanism***: a patient may move into a faster speed category without meaningful normalization of COM work organization. Practically, this means that speed can improve through further compensation, i.e. greater reliance on the non-paretic limb and altered work distribution and may therefore overstate paretic limb mechanical recovery if interpreted alone. This aligns with contemporary perspectives arguing for propulsion-focused diagnostics and treatment development, precisely because propulsion and its timing reflect limb-level function beyond global performance metrics^23^.

The present analysis is speed-stratified and does not establish causality between COM work pattern restoration and patient-centered outcomes. Nevertheless, the observed nonlinearity and piecewise behavior across indices support the concept that hemiparetic walking traverses distinct mechanical regimes as speed changes. A logical next step is prospective validation: testing whether COM work/PSR metrics predict clinically meaningful endpoints e.g., endurance, real-world mobility, falls risk, beyond gait speed, consistent with the field’s increasing emphasis on pairing clinical outcomes with more specific mechanistic phenotyping ^24^. It also has the potential to distinguish between compensatory strategies and recovery-based restoration.

The distribution of COM work patterns (Figure 3) together with the derived indices may provide clinicians with a biomechanically grounded framework to stratify hemiparetic gait severity and to objectively monitor rehabilitation progress. In particular, the absence of paretic push-off, reflected by a two-phase COM power trajectory consisting only of net positive and negative work, appears to characterize a mechanically severe state. Conversely, emergence of paretic limb push-off and restoration of a recognizable four-phase COM power profile indicates partial reorganization of transition mechanics and may correspond to a milder deficit. Importantly, when paretic push-off is present but the unaffected limb’s preload (single-support negative work) remains minimal, this pattern may represent an intermediate state—suggesting transition from severe to less severe mechanical impairment. The presence of this transitional state is supported quantitatively by the observed quadratic turning points and piecewise breakpoints in propulsion asymmetry and PSR trends, which cluster around the same speed domain.

Among the evaluated metrics, the Propulsion–Support Ratio (PSR), considered jointly for paretic and unaffected limbs, appears particularly robust for severity characterization because it directly reflects the limb’s ability to convert stance support into forward impulse. When interpreted along with the COM work distribution map, PSR provides a concise yet mechanistically informative descriptor of paretic gait severity that may complement traditional asymmetry indices such as AI_W_net_ and AI_W_pos_.

These findings support a pragmatic refinement—not a replacement—of current severity assessment: use gait speed as the functional headline but supplement it with a small set of COM work and propulsion–support indices. Speed remains essential for clinical benchmarking and communication, but COM work organization can reveal whether a faster gait reflects mechanical restoration of paretic limb function or continued reliance on compensatory work redistribution. This distinction is actionable: it can guide selection and evaluation of propulsion-and loading-targeted interventions, reduce the risk of misclassifying compensated walking as recovery, and identify biomechanical thresholds where meaningful reorganization of transition mechanics emerge. In short, pairing speed with COM-based mechanistic metrics offers a feasible path toward mechanism-informed stroke rehabilitation while preserving the interpretability and clinical value of conventional performance measures ^3^.

## Data Availability

The data used in these analyses are available at the attached repository

https://github.com/salehhosseini/Post-Stroke-HemiParetic-Walking

## Acknowledgment

This work was supported by a Natural Sciences and Engineering Research Council of Canada (NSERC) Discovery grant (04823-2017) received by J.E.A.B.

## Notes

**Conflict of Interest:** None

### Competing Interest Statement

The authors have declared no competing interest.

### Funding Statement

This work was funded in part by NSERC Discovery grant 04823-2017 (to JEAB) and through a donation from the Rodie Foundation

### Author Declarations

Written informed consent obtained according to a protocol approved by the Conjoint Health Research Ethics Board at the University of Calgary (Ethics ID REB21-1576).

